# Physician gestalt compared with AI model to predict intubation in critically ill patients

**DOI:** 10.64898/2025.12.19.25342667

**Authors:** Michael A Miller, Xiaolei Lu, Alex Pearce, Atul Malhotra, Shamim Nemati

## Abstract

**Rationale:** Intubation and mechanical ventilation are associated with high mortality. Accurately predicting which patients are at the highest risk of intubation can enable interventions to reduce their risk. The performance of intensive care physicians to predict the need for intubation within the next 24 hours for medically critically ill patients is unknown. Machine learning models are adept at prediction tasks.

**Objective:** In this study, we perform a prospective observational study in two ICUs to survey intensivists to test their accuracy at predicting the need for intubation within 24 hours of patients under their care. Physician predictions of intubation are then compared to predictions from a machine learning model called Vent.io.

**Methods:** Primary metrics included prediction sensitivity, specificity, and descriptive statistics for both physician and machine learning model. Generalized linear mixed models were developed to investigate the fixed effect of the predictor (physician vs Vent.io) on both sensitivity and specificity while accounting for the random effects from different physicians and reported by odds ratio and 95% confidence interval. Similar modeling was also used to test the relationship between physician confidence and correctness.

**Results:** Overall, physicians are quite confident in their predictions of intubation with a median score of 8 (on a 0–10 point scale, with 0 being not at all confident and 10 being extremely confident) out of the 302 surveys administered. Sensitivity was 0.190 and 0.714 for physicians and Vent.io, respectively. Specificity was 0.960 and 0.673 for physicians and Vent.io, respectively. Generalized linear mixed modeling showed that physician confidence is associated with greater odds of correctly predicting intubation outcome (OR 1.49; 95% CI 1.22-1.84; p<.001). Vent.io had significantly greater odds of being correct when patients required intubation compared to physicians (OR 18.68; 95% CI 1.87-186.31; p=0.013). However, intensive care physicians outperformed Vent.io at correctly predicting when patients did not require intubation (OR 24.80; 95% CI 13.22-46.52; p<0.001).

**Conclusions:** While promising, Vent.io needs real-time testing in a randomized clinical trial to determine if its deployment can improve clinical outcomes.

## Introduction

Acute respiratory failure (ARF) is a common condition in the intensive care unit (ICU).^1^ In addition to treating the underlying cause of ARF, physicians often need to support critically ill patients with intubation and mechanical ventilation. Emergent intubation is a high-risk procedure.^2^ Despite advances in the field, invasive mechanical ventilation (IMV) for ARF is associated with high mortality,^3^ and interventions aimed at decreasing need for intubation must first have an accurate estimate of which patients with ARF will require the procedure.^4^ Ideal prediction models provide enough runway for clinicians to perform interventions that can either avoid the need for mechanical ventilation altogether or increase patient safety by avoiding complications inherent with late identification of severe progressive ARF requiring IMV.

The optimal timing of intubation in ARF is uncertain and depends on weighing the risks and benefits of initiating IMV.^5^ Clinical decision-making for intubation timing is complex, and clear clinical practice guidelines from relevant medical societies are lacking. Knowledge about how well clinicians predict the need for intubation in advance for medically critically ill patients is also lacking. In recent years, many predictive analytic models utilizing machine learning have arisen to predict outcomes in the acute respiratory failure space, including the advanced prediction of intubation.^6–8^

The present study has two main objectives. The first is to understand how well intensive care physicians predict the need for IMV within 24 hours. After collecting binary predictions from physicians, their confidence, and ground truth outcomes, we will describe their performance metrics. The second objective is to compare the performance of intensive care physicians versus an established machine learning model to predict whether a critically ill patient will require IMV in the next 24 hours. We hypothesize that the machine learning model will have higher sensitivity but lower specificity for predicting IMV compared with the physicians at this task.

## Methods

We conducted a prospective, longitudinal, observational study from October 2024 until May 2025. Intensivist attendings at two medical ICUs at two University of California San Diego (UCSD) hospitals participated. These closed teaching ICUs are within academic tertiary care centers and predominantly treat patients with acute respiratory failure, kidney failure, sepsis, liver failure, and cancer complications. Ethics board approval was obtained (UCSD IRB #811257).

### Population

All intensivists caring for patients in the medical ICU during the study period were invited to participate. On a daily basis, when available, a pulmonary and critical care fellow screened the ICU lists to identify non-intubated adult patients admitted to the ICU service without limited code status (do not intubate order) or tracheostomy. Attending intensivists were approached (mostly in person but sometimes by phone call) to ask if they would be willing to participate in a brief survey. The survey collected predictions of whether the patients that the physicians were taking care of (who were identified by the screening process) would require intubation and mechanical ventilation within the next 24 hours.

### Intensivist Survey

A survey was used to collect physician predictions for the outcome of invasive mechanical ventilation within 24 hours. The survey was brief and designed to be completed in under two minutes to avoid clinical workflow interruptions. The first question was binary: “Do you think *Patient X* will require invasive mechanical ventilation or not within the next 24 hours?” Next, the intensivists were asked to provide confidence in their answer from 0-10, where 0 represented “not at all confident in my answer” and 10 represented “extremely confident in my answer.” The date and time were recorded along with a patient identifier. ICU physician experience metrics (number of years since completing training and number of weeks worked in the ICU per year) were also collected. They were also asked whether a dashboard, integrated into the electronic health record, that predicted whether a patient was at high risk of intubation in the next 24 hours, would be helpful or not. All data were recorded into a REDCap database. Patients requiring intubation for planned procedures (e.g., liver transplant) were excluded to avoid capturing patients intubation for airway protection during anesthesia instead of intubation for clinical deterioration. A total of 300 surveys was selected for the enrollment target as that was the number of surveys thought to be feasible to obtain during the time allotted for the study.

### AI prediction

A previously developed machine learning (ML) model called Vent.io^6^ was used to predict the need for IMV within 24 hours. The model architecture is a 3-layer feedforward artificial neural network trained on hourly binned electronic health record (EHR) data (vital signs, labs, demographics, SIRS/SOFA criteria, medications, and comorbidities) from adult ICU patient encounters and had median AUC 0.897 and 0.908 during cross validation and prospective deployment, respectively.^6^ The model’s output is a numeric score that reflects risk of intubation within the next 24 hours, and a preset decision threshold corresponding to 60% sensitivity was used to convert to a binary prediction of “yes” or “no.” The model was queried at the same timestamp that the intensivists were surveyed to ensure that both physician and model predictions had the same information contemporaneously available to them at the time of prediction. Both the ML model and the physicians were blinded to each other’s predictions.

### Outcomes and Analysis

After predictions were obtained from the physicians and Vent.io, patients were then followed to obtain the ground truth outcome (i.e. did intubation occur within the subsequent 24 hours). Descriptive statistics were used to describe the surveyed population, occurrence of intubation, and data missingness. The primary outcomes for this study were the sensitivity and specificity for each prediction method (physician and Vent.io). Generalized linear mixed models were developed to investigate the fixed effect of the predictor (Vent.io vs physician) on both sensitivity and specificity while accounting for the random effects from different physicians making predictions. Confidence intervals (CIs) were obtained by bootstrapping.

As a secondary outcome, the composite metric of balanced accuracy of each prediction method (physicians as a whole, not adjusted for the random effects of multiple physicians, and the ML model) was reported with 95% CIs. Balanced accuracy was selected given its interpretability and the low incidence of intubation. The performance of Vent.io was also described by area under the receiver operator curve (AUC), and optimal thresholding for this dataset was investigated with Youden’s statistics. Another generalized linear mixed methods model was developed to predict physician correctness based on physician confidence. The model accounted for the clustering of observations by physicians and included terms to test if the relationship between confidence and correctness was moderated by the physician experience metrics of years since training and weeks worked per year in the ICU. Classification consistency between physician and Vent.io predictions was also compared with McNemar’s Test. The analysis was performed using R, version 4.4.3, and included the packages *lme4, caret, boot*, and *dplyr*.

## Results

### Descriptive Statistics

A total of 302 surveys were completed over the study period. Intubation within 24 hours of prediction occurred in 21 (7.0%) of those instances. The amount of time needed to complete the surveys was estimated at less than two minutes per attending per day surveyed. There was no missingness for physician predictions of 24-hour need for intubation, physician confidence, Vent.io predictions, or ground truth outcomes. Twenty-four board certified intensive care physicians participated during the study period. The group was 21% female, worked a median of 10 weeks on service in the ICU per year, and was a median of 11 years post fellowship training.

### Physician Performance

The physicians predicted 15 (5.0%) times that patients would require intubation within the subsequent 24 hours and 287 (95%) times that they would not require intubation. Overall, the intensivists correctly predicted intubation for 4 of the 21 intubated patients (sensitivity 0.190) and correctly predicted no intubation for 270 of the 281 instances in which patients were not intubated in the 24 hours following the time of survey administration (specificity 0.960). The balanced accuracy of physicians (without accounting for the random effect of different physicians) was 0.576 (95% CI 0.499 - 0.666). The median confidence in their prediction was 8 (on a 0-10 scale, with 0 being not at all confident and 10 being extremely confident) [mean 7.4, SD 2.2][Figure 1]. The generalized linear mixed methods model evaluating how physician confidence predicts correctness revealed that higher physician confidence is associated with greater odds of being correct (OR 1.49; 95% CI 1.22-1.84; p<.001). No significant influence from physician experience modifiers was identified.

**Figure 1.**
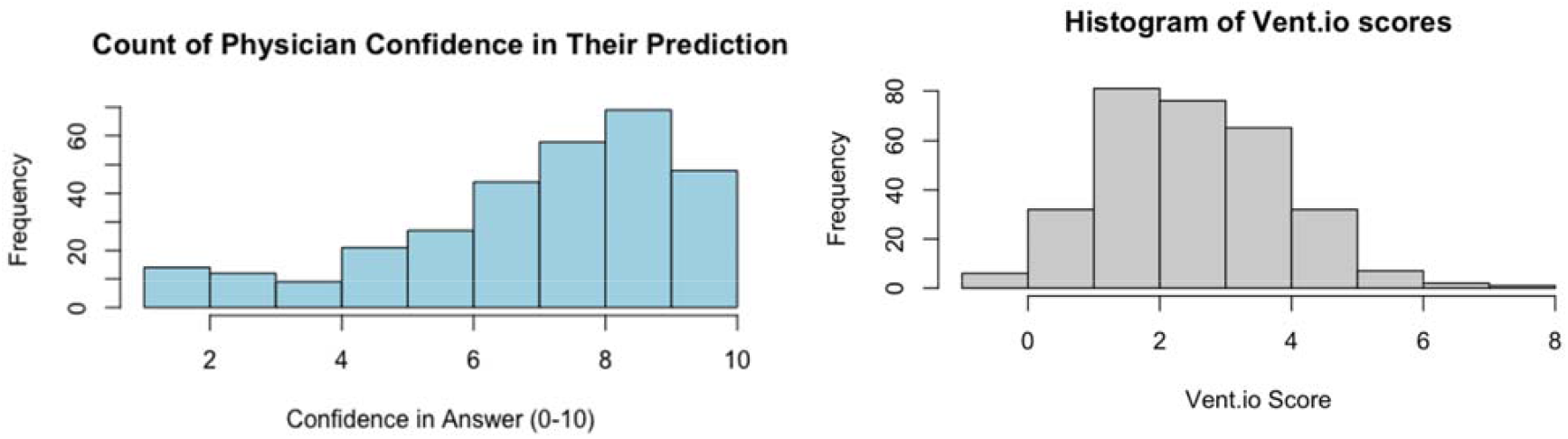
On the **left** is a histogram depicting the spread in confidence levels of physician for their predictions. On the **right** is a similar histogram that shows the frequency of Vent.io scores from the ML model. Note that a Vent.io score of ≥3 is used for a model prediction of “yes” for intubation.

### ML model performance

The Vent.io model outputs ranged from −0.71 to 7.21 with a mean value of 2.51 and a standard deviation of 1.34. Predictions (above the set threshold of 3) for intubation within the next 24 hours numbered 107 (35.4%) and predictions for no intubation numbered 195 (64.6%). Overall, Vent.io correctly predicted 15 of the 21 patients who were intubated (sensitivity 0.714), and Vent.io correctly predicted no intubation in 189 of the 281 instances in which patients were not intubated (specificity 0.673) in the subsequent 24 hours (Figure 2). Compared to the physicians, the Vent.io model had significantly greater odds of a correct prediction when the patient did require intubation (OR 18.68; 95% CI 1.87-186.3; p=0.013). Conversely, the average physician had significantly greater odds of correctly predicting that a patient would not require intubation in the next 24 hours compared with Vent.io (OR 24.80; 95% CI 13.22-46.52; p<0.001). The balanced accuracy of Vent.io was 0.693 (95% CI 0.587-0.794). The area under the receiver operating curve (AUC) for Vent.io was 0.798 (95% CI 0.717-0.972). The optimal Vent.io decision threshold (by Youden’s J) for this dataset of 2.365 was lower than the set threshold of 3. Exact McNemar’s Test (chi-squared = 82.8; p<0.001) showed there was statistically significant difference in the error patterns of the two methods such that the disagreements are more likely in the direction of Vent.io predicting “yes” and the physicians predicting “no.”

**Figure 2.**
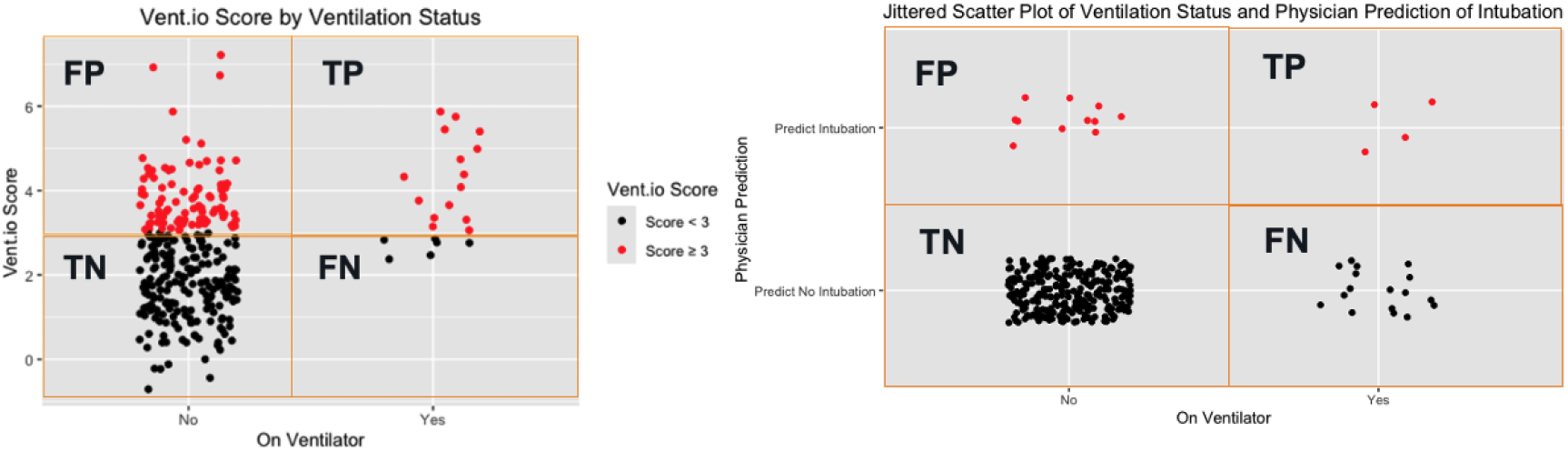
On the **left** is a jittered scatterplot demonstrating Vent.io score in the conditions that patients were or were not intubated within 24 hours. A red dot represents a prediction for intubation, and each black dot represents a prediction of no intubation. On the **right** is a jittered scatterplot where the x-axis represents the two ground truth outcomes of intubated or not intubated, and the y-axis represents a physician prediction of “the patient will need intubation within 24 hours (red)” or “the patient will not need intubation (black).” Together these scatterplots visually represent the confusion matrices used to calculate the main performance metrics.

At the end of the study period, 18 of the 24 (75%) physicians answered the question, “If you had access to ML tool in a dashboard [in the electronic health record] that could accurately predict whether a patient would require intubation in the next 24 hours, would you find that useful or not?” Thirteen (72%) thought this would be a useful feature to have.

## Discussion

In this prospective cohort study, which examined the ability of both intensive care physicians and a machine learning model to predict the 24 hour need for intubation in critically ill patients, there were several interesting findings. First, despite high confidence in their ability, performance at this task was quite poor; only 19% of patients who ultimately required intubation were correctly identified by the physicians ahead of time. The ML model significantly outperformed physicians at this task. At the task of correctly identifying those medical ICU patients who will not require intubation, physicians did much better, outperforming the ML model with a specificity of 96%.

To the best of our knowledge, this paper describes the first assessment of intensive care physicians’ ability to predict the advanced intubation needs of medically critically ill adults. Additionally, we know of no prior examples of head-to-head assessments of physicians versus a machine learning model of performance at this task. Predicting the 24 hour need for intubation is akin to a screening task. In this context, higher sensitivity than specificity might be preferred to avoid the risk of missing positive cases (false negatives). When intensivist sensitivity is lower, patients who require intubation are identified later. Clinically this means that interventions that require time for effect (antibiotics, steroids, changing respiratory support modality, goals of care discussions, etc.) might happen later, which could lead to worse clinical outcomes.

If a model like Vent.io were ultimately embedded into the EHR for physicians to review (this capability already exists^6^), higher sensitivity might be valuable. Physicians could use such a dashboard when less familiar with the patients (cross-covering overnight, first day on service, etc.) or as a prompt to reassess a patient if there is a substantial worsening in their Vent.io score. Anecdotally, none of the physicians surveyed anticipated ever intubating a patient solely because of a high Vent.io score, but many said that a concerning score would lead them to reevaluate the patient. If this approach is how clinicians interact with the tool, then the risk to the patients of a false positive score is low and the primary risks of false positives are to physicians’ time (reevaluating a patient who did not require reevaluation) or alarm fatigue. Physicians behaved in a way that maximized specificity at the expense of sensitivity most likely to avoid the situations where a patient is intubated who does not truly require it.

This study had several strengths and weaknesses. A broad group of intensivists (over 20) participated, and data missingness was minimal. The primary outcomes were appropriate and helped answer the main clinical questions of the study. Vent.io predictions came in a continuous format which was then transformed to a binary “yes/no” prediction, while clinician predictions were binary and then prediction confidence was collected. Comparison would have been more straightforward if physicians were asked to predict the likelihood of intubation in the next 24 hours. The incidence of intubation was low, occurring only 21 times out of 302 prediction instances, which may have caused some selection bias. This result is likely because some patients were intubated in the emergency department, wards, or in the ICU overnight before a survey was administered during the daytime in the ICU. While the directionality of the ORs is plausible, the magnitude is possibly overstated. This is evidenced by the wide confidence intervals. There is likely imprecision in the OR estimates given the rarity of intubation (7%, which introduces the potential for sampling error) in a smaller sample size (302 surveys).

The training of the Vent.io model included patients in medical ICUs but also included patients from other ICU types. The lower optimal Vent.io decision threshold by Youden’s statistic for our dataset could reflect either the different populations or be related to the possible sampling error discussed above. Another interpretation of the limits to the sensitivity and specificity of both the physicians and Vent.io is that predicting intubation 24 hours in advance in a medical ICU population is quite a difficult task regardless of the prediction technique, stemming from the complicated clinical picture of many of these patients, as well as the quick pace at which clinical changes and medical interventions occur in this population.

Despite Vent.io’s successful performance in the early identification of medical ICU patients at high risk for intubation, more steps are needed before this model can be used in clinical practice. The understanding of model performance relative to physician performance in this study is important for framing the contexts in which Vent.io might be most useful clinically. For example, knowing physician sensitivity is low, the model could act as a flag or early warning sign that the patient is at elevated risk of intubation and prompt the care team to consider reassessment or evaluate whether a checklist of respiratory interventions would be appropriate for the patient. Before Vent.io can be utilized as a decision support tool in hospitals, it has to be more rigorously tested to show that it improves clinically meaningful outcomes. The model’s current ability to deploy in the EHR in real-time and randomize to “live” or “silent” mode (from the perspective of the clinical care team) should make a randomized clinical trial feasible. Further investigation can explore the patient level characteristics associated with either correct or incorrect predictions of intubation by both the ML model and the clinicians.

## Conclusion

In conclusion, this study shows that, despite having high confidence in their ability to predict whether critically ill patients will require intubation in the next 24 hours, physicians are poor predictors when patients do require intubation but good predictors when patients do not require intubation. Compared to a ML model called Vent.io which performs the same prediction task, physicians have significantly worse sensitivity but better specificity. The predictive algorithm should be tested in a randomized control trial to evaluate whether its deployment improves clinical outcomes or not. Ultimately combining physician judgment with machine learning techniques may be necessary to optimize clinical outcomes.

## Data Availability

All data produced in the present study are available upon reasonable request to the authors

## Financial/non-financial disclosures

This work was supported by the National Heart, Lung, and Blood Institute (R01HL157985). S.N. and A.M. have equity and are cofounders of a UC San Diego start-up, Clairyon Inc. (formerly Healcisio Inc.), which is focused on the commercialization of advanced analytical decision support tools, and formed in compliance with UC San Diego conflict of interest policies. A.M. is funded by the NIH. He reports income from Zoll, Livanova, Eli Lilly, Powell Mansfield and Sunrise. Resmed gave a philanthropic donation to UCSD.

## Other disclosures

Miller: None

Lu: none

Pearce: none

## Role of sponsors

The funders had no role in the study.

## Acknowledgement of contributions

MM designed the study, collected the data, performed the statistical analyses, interpreted the results, compiled the manuscript and is accountable for all aspects of the work. XL and SN developed the ML model and collected model outputs, interpreted the results, and provided critical revisions for the manuscript. AP interpreted the results and provided critical revisions for the manuscript. AM and SM consulted on the study design, refined the analyses, assisted in interpreting the findings, and provided critical revisions of the manuscript.

## Abbreviations

ARF: acute respiratory failure
ICU: intensive care unit
ML: machine learning
AI: artificial intelligence
AHRF: acute hypoxic respiratory failure
OR: odds ratio
CI: confidence interval
AUC: area under the receiver operator curve
IMV: invasive mechanical ventilation
EHR: electronic health record
TP: true positive
TN: true negative
FP: false positive
FN: false negative

## AI Disclaimer

No AI tools were used to write this manuscript

## Acknowledgments

The authors thank Hayden Pour for his work in the lab of Shamim Nemati that supports research efforts like this.

## Notes

### Author Declarations

Ethics board approval was obtained (University of California San Diego's Institutional Review Board #811257).

## Citations

1. Villar J, Mora-Ordoñez JM, Soler JA, et al. The PANDORA study: Prevalence and outcome of acute hypoxemic respiratory failure in the pre-COVID-19 era. Crit Care Explor. 2022;4(5):e0684.

2. Russotto V, Myatra SN, Laffey JG, et al. Intubation practices and adverse Peri-intubation events in Critically Ill Patients from 29 countries. JAMA. 2021;325(12):1164–1172.

3. Bellani G, Laffey JG, Pham T, et al. Epidemiology, patterns of care, and mortality for patients with acute respiratory distress syndrome in intensive care units in 50 countries. JAMA. 2016;315(8):788–800.

4. Pearce AK, Nemati S, Goligher EC, et al. Can we predict the future of respiratory failure prediction? Crit Care. 2025;29(1):253.

5. Lee KG, Roca O, Casey JD, et al. When to intubate in acute hypoxaemic respiratory failure? Options and opportunities for evidence-informed decision making in the intensive care unit. Lancet Respir Med. 2024;12(8):642–654.

6. Lam JY, Lu X, Shashikumar SP, et al. Development, deployment, and continuous monitoring of a machine learning model to predict respiratory failure in critically ill patients. JAMIA Open. 2024;7(4):ooae141.

7. Liu J, Duan X, Duan M, et al. Development and external validation of an interpretable machine learning model for the prediction of intubation in the intensive care unit. Sci Rep. 2024;14(1):27174.

8. Venturini M, Van Keilegom I, De Corte W, Vens C. Predicting time-to-intubation after critical care admission using machine learning and cured fraction information. Artif Intell Med. 2024;150(102817):102817.

